# A methodological framework to assess temporal trends and sub-national disparities in healthcare quality metrics using facility surveys, with applications to sick-child care in Kenya, Senegal, and Tanzania

**DOI:** 10.1101/2022.07.19.22276796

**Authors:** Adrien Allorant, Nancy Fullman, Hannah H. Leslie, Moussa Sarr, Daouda Malick Gueye, Eliudi Eliakimu, Jonathan Wakefield, Joseph L. Dieleman, David Pigott, Nancy Puttkammer, Robert C. Reiner

## Abstract

Monitoring healthcare quality at a subnational resolution is key to identify and resolve geographic inequities and ensure that no sub-population is left behind. Yet, health facility surveys are typically not powered to report reliable estimates at a subnational scale.

In this study, we present a framework to fill this gap and jointly analyse publicly available facility survey data, allowing exploration of temporal trends and subnational disparities in healthcare quality metrics. Specifically, our Bayesian hierarchical model includes random effects to account for differences between survey instruments; space-time processes to leverage correlations in space and time; and covariates to incorporate auxiliary information. We apply this framework to Kenya, Senegal, and Tanzania - three countries with at least four rounds of standardized facility surveys each – and estimate the readiness and process quality of sick-child care over time and across subnational areas.

These estimates of readiness and process quality of care over time and at a fine spatial resolution show uneven progress in improving facility-based service provision in Kenya, Senegal, and Tanzania. For instance, while national gains in overall readiness of care improved in Tanzania, geographic inequities persisted; in contrast, Senegal, and Kenya experienced stagnation in overall readiness at the national level, but disparities grew across subnational areas. Overall, providers adhered to about one-third of the clinical guidelines for managing sick-child illnesses at the national level. Yet across subnational units, such adherence greatly varied (e.g., 25% to 85% between counties of Kenya in 2020).

Our new approach enables identifies precise estimation of changes in the spatial distribution of healthcare quality metrics over time, at a a programmatic spatial resolution, and with accompanying uncertainty estimates. Use of our framework will provide new insights at a policy-relevant spatial resolution for national and regional decision-makers, and international funders.

## Introduction

Making progress toward universal health coverage (UHC) is recognised as a central strategy to improve equity in health service delivery, and thus achieve better health outcomes for all ^1^. Much debate remains around the ideal measurement of UHC advances, with the increasing recognition that monitoring UHC will involve substantially strengthening nationally representative and timely data systems to optimally measure access and the provision of high-quality healthcare ^2^. In their absence, health facility surveys, including the Service Provision Assessment (SPA) and the Service Delivery Indicators (SDI), remain primary data sources for assessing and comparing the quality of available health services in low- and middle-income countries (LMICs) ^3–8^. These facility surveys provide detailed information about service components and capacities, as grouped into structures (basic amenities, infection control, equipment, diagnostics, and medication), processes (components of clinical care), and outcomes, including patients’ satisfaction with services received ^9^.

Recent studies using SPA and SDI surveys have operationalized assessment guidelines or clinical charts provided by the World Health Organization (WHO) to describe countries’ service availability, readiness, and process quality of care ^5,10–13^. Yet, most analyses to date are restricted to the most recent survey and focus on cross-sectional analyses to provide national-level estimates of healthcare availability, readiness, and quality. Novel research has compared country-level estimates from several years of facility surveys ^14–16^, but differences in sample size and design can limit their actual comparability. Statistical modelling frameworks offer means by which such differences can be explicitly accounted for and support trend estimation in readiness and process quality indicators over time. Additionally, recent work has highlighted large subnational disparities in the coverage of key health services ^17^, interventions ^18^ and outcomes ^19^, in sub-Saharan African countries, underscoring the need to better understand and monitor sub-national variations in health system inputs and processes ^20^. The decentralization of health services in many sub-Saharan countries has substantially increased local governments’ responsibilities in the planning and implementation of public health ^21–23^, including the maintenance and equipment of health facilities, which has generated an enhanced demand for sub-national indicators ^24^.

In this study, we present a statistical framework to assess changes in the quality of facility-based health services at a subnational scale, despite data challenges associated with size, survey design, and comparability. Our model supports the use of publicly available facility data in each country while accounting for the differences between survey instruments, and supplements direct survey measurements with auxiliary information from spatiotemporally indexed covariates. We demonstrate the application of our approach by estimating metrics of readiness and process quality of sick-child care in Kenya, Senegal, and Tanzania, over time and across subnational areas.

## Methods

### Data sources

Facility data come from two standardized health facility assessment tools that are publicly available and collect information on process quality: the SPA and the SDI. The SPA and SDI are consistent and comparable health facility surveys, designed to be nationally representative of the formal health sector. SPA surveys generally use a stratified survey design by facility type (e.g., hospital, health centres, clinics), managing authority (e.g., public/private), and first administrative division, and typically include four modules: an inventory questionnaire, observations of consultations, exit interviews with the observed patients, and interviews with healthcare providers (https://dhsprogram.com/methodology/Survey-Types/SPA.cfm/). The SDI survey is stratified by urban/rural areas and first administrative division, and comprises three modules: an inventory questionnaire, clinical vignettes to assess providers’ knowledge, and unannounced visits to facilities to measure providers’ absenteeism (https://www.sdindicators.org/).

In this analysis, we used all publicly available cross-sectional facility surveys in Kenya, Senegal, and Tanzania, three countries with at least four facility surveys each, conducted from 1999 to 2019, and thus supported estimating levels and trends in readiness and process quality of care across subnational areas. We focused on child health services as they are the targets of large national and international investments in all three countries ^25^. We analysed data from 10,431 facilities offering child curative care, 18,693 direct observations of sick-child consultations and 6,283 clinical vignettes, in Kenya (1999-2018), Senegal (2012-2019), and Tanzania (2006-2016) (see Table 1) to characterize facility-level information relating to the availability, readiness and process quality of child health. Additionally, we used facilities’ sampling weights, and the survey design variables – facilities’ first administrative location, type and managing authority.

**Table 1:**
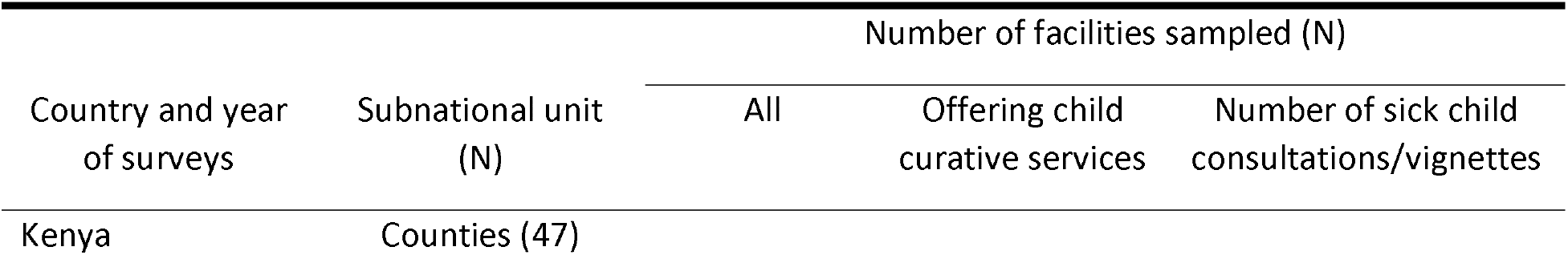
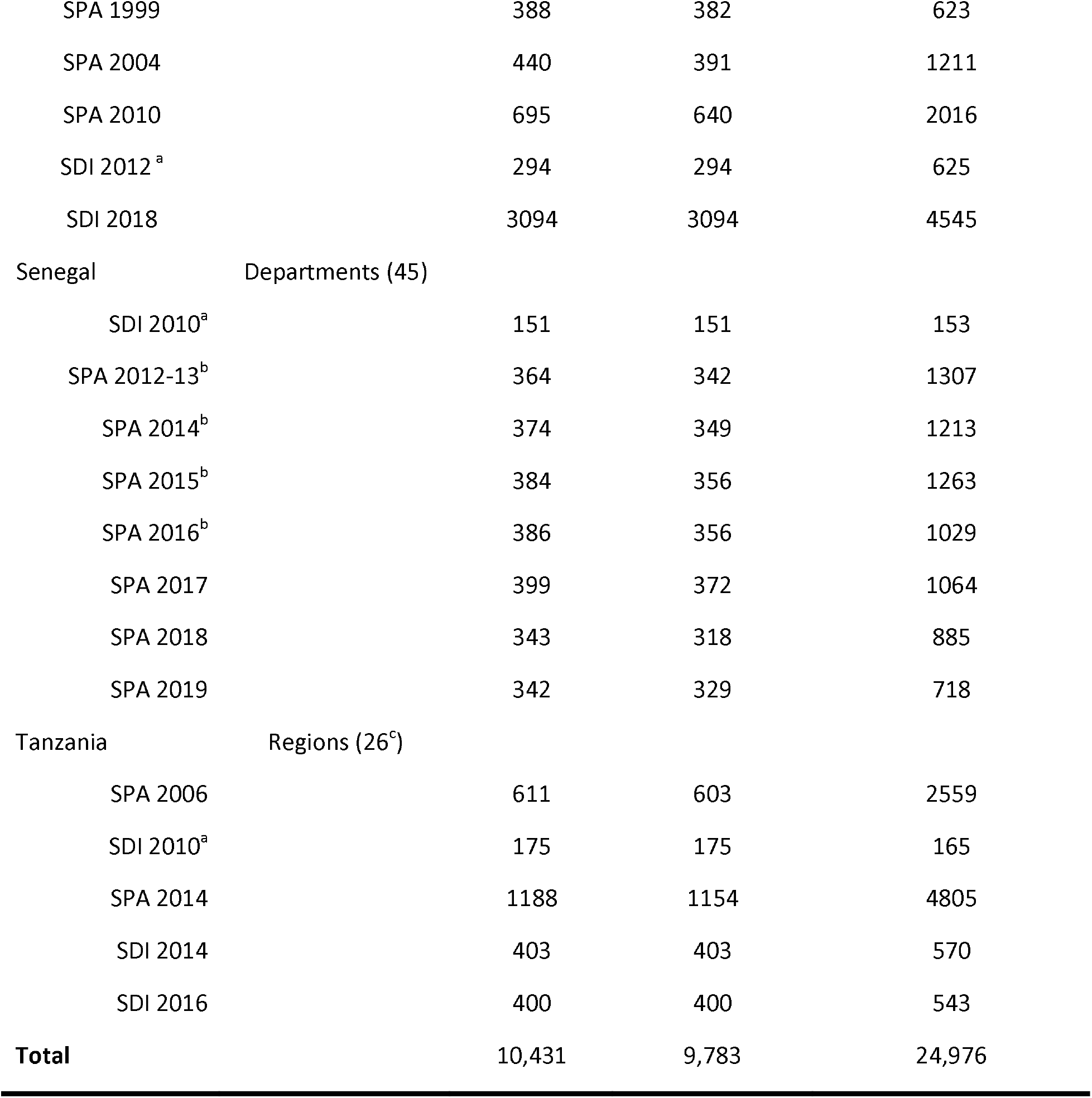
List of surveys publicly available in Kenya, Senegal, and Tanzania and their sample size. ^a^ The 2010 SDI surveys in Senegal and Tanzania and the 2012 SDI survey in Kenya were pilot studies, which only sampled a small number of facilities in selected areas of the countries and were therefore excluded from the main analysis. ^b^ Dependent sampling structure between the first four rounds of the continuous SPA SPA-survey in Senegal 2013-2016 (see Supplementary Table S1). Data from SPA 2012-13 and 2014, and from 2015 and 2016 were pooled to mitigate the effect of the dependent sampling on estimates’ comparability. ^c^ Tanzania was divided into twenty-six regions at the beginning of our study period in 2006. Although four new regions were created in 2012, and one in 2016 (such that Tanzania now counts 31 regions), we used the twenty-six-region divide in our analyses to ensure that our results would not be affected by historical boundary changes.

### Measures

We extracted items reflective of general and child curative services readiness listed in the WHO SARA framework ^26^ and processes of care listed in the Integrated Management of Childhood Illness (IMCI) ^27^. The readiness metric was based on the availability and functioning of general equipment, basic amenities, staff training and supervision, essential medicines, and diagnostic capacities, specific to the provision of child curative care services. The items included in the readiness metric were selected based upon previous guidelines and research in paediatric quality of care ^27,28^. As some of these items were not collected in the SDI surveys and older SPA surveys, we modified the two indices in Kenya and Tanzania (staff training and supervision items were excluded) to ensure comparability of readiness estimates across years in each country (see Supplementary Table S4). The readiness metric was calculated, for each health facility offering child curative care, as the proportion of items available.

We derived the metric of process quality of care from the content of sick child consultations observed in the SPA surveys, and the clinical knowledge displayed by health providers sampled for vignettes in the SDI surveys. Following previous studies ^7,29^, adherence to IMCI diagnostic protocols was used as a proxy for process quality of sick-child care. Our metric of process quality of care was calculated as the proportion of fifteen IMCI diagnostic protocols adhered to by providers during sick-child consultations or vignettes (see Supplementary Table S5).

### Statistical Analyses

Facility survey data were complemented with covariates, which have a known or postulated relationship with health services provision. The data set of temporally varying covariates at the subnational level included total population under five years old, travel time to nearest settlement of more than 50,000 inhabitants, travel time to nearest health facility (walking and motorized), health worker density, urbanicity, night-time lights, average educational attainment, human development index, and elevation. We extracted secondary data to complement facility survey data, using sources like WorldPop or IHME, to identify pertinent indicators (see complete list of data sources and processing in Supplementary Table S3).

The metrics of readiness and process quality of care were modelled separately using a previously-developed small area estimation approach ^30,31^. First, facility-level metrics of readiness and process quality were aggregated at the subnational area-level using facilities’ sampling weights. Explicitly incorporating surveys’ sampling mechanism through the use of weights is a common method for bias removal when analysing complex surveys ^32–34^. Second, we estimated seven multi-level logistic models, which represented subnational area-level readiness and process quality metrics as a function of both independent and temporally structured year random effects (model 1-7), independent (model 1-4) or spatially correlated areal random effects (model 5-7), spatially and temporally indexed covariates (listed in Supplementary Table S3) that we hypothesised to be predictive of health service provision (model 3,4 and 7), and an indicator variable identifying survey type (model 2,4,6 and 7; see Supplementary Section 2.2-2.5 for more details on the models). While space-time random effects and covariates were used to improve the precision of our estimates by leveraging correlation structures in space, time and with auxiliary variables, survey indicators were utilized to account for systematic differences (in design or implementation) between survey instruments. For each metric, we used goodness of fit and model complexity to identify the best performing model out of the seven models (Supplementary Section 2.5). We then generated and mapped annual area-level estimates of the two metrics for each Kenyan county from 1999 to 2020, each Senegalese department from 2010 to 2020, and each region of Tanzania from 2005 to 2020. Estimates and their associated 95% uncertainty intervals were obtained by drawing 1,000 posterior samples for all parameters estimated in the model and calculating the mean, and the 2.5 and 97.5 quantiles. We used these estimates to characterize changes in quality over time and to quantify levels of inequities between subnational areas.

To test the predictive validity of our models, we removed all observations for a given area and year and refit each model holding out those observations. We then compared the areal model’s prediction with the observed direct estimate derived from the survey. This was completed for all the areas and years for which we survey data were available. For each metric, we assessed the quality of our predictions by examining the average distance between the predictions and the observed estimates (mean squared error - MSE), and by calculating the 80% coverage, i.e., the frequency at which the direct survey estimate was contained within the model’s predicted 80% uncertainty interval.

## Results

### Survey estimates of readiness and process quality metrics

Average facility readiness was stable over the study period in Kenya and Senegal, around 60-65% nationally; in contrast, Tanzania showed marked improvements, from 54.4% (53.6-55.3) in 2006 to 74.6% (72.6-76.6) in 2016 (Table 2). For readiness of care, the main constraints to higher performance were related to facility management items (quality assurance, recent supervision) and essential drug stocking (amoxicillin and co-trimoxazole for children), as well as malaria diagnostics, especially in Kenya and Tanzania (Supplementary Table S7-S9 provide item availability by country and survey). Process quality of care had steady improvements in Kenya, from 30.7% (29.7-31.7) in 1999 to 43.7% (42.7-44.7) in 2018, and stagnation in Senegal and Tanzania around 35-40% over the survey time periods.

**Table 2:**
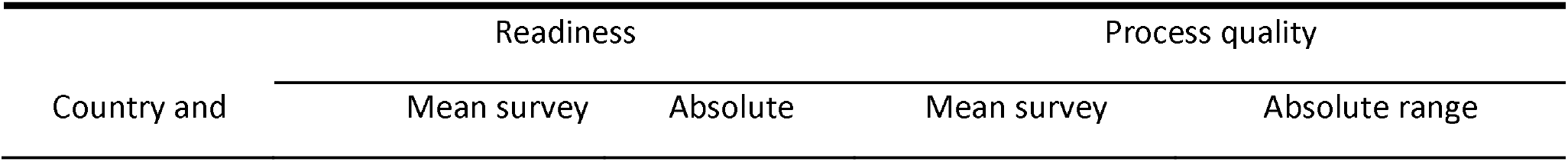
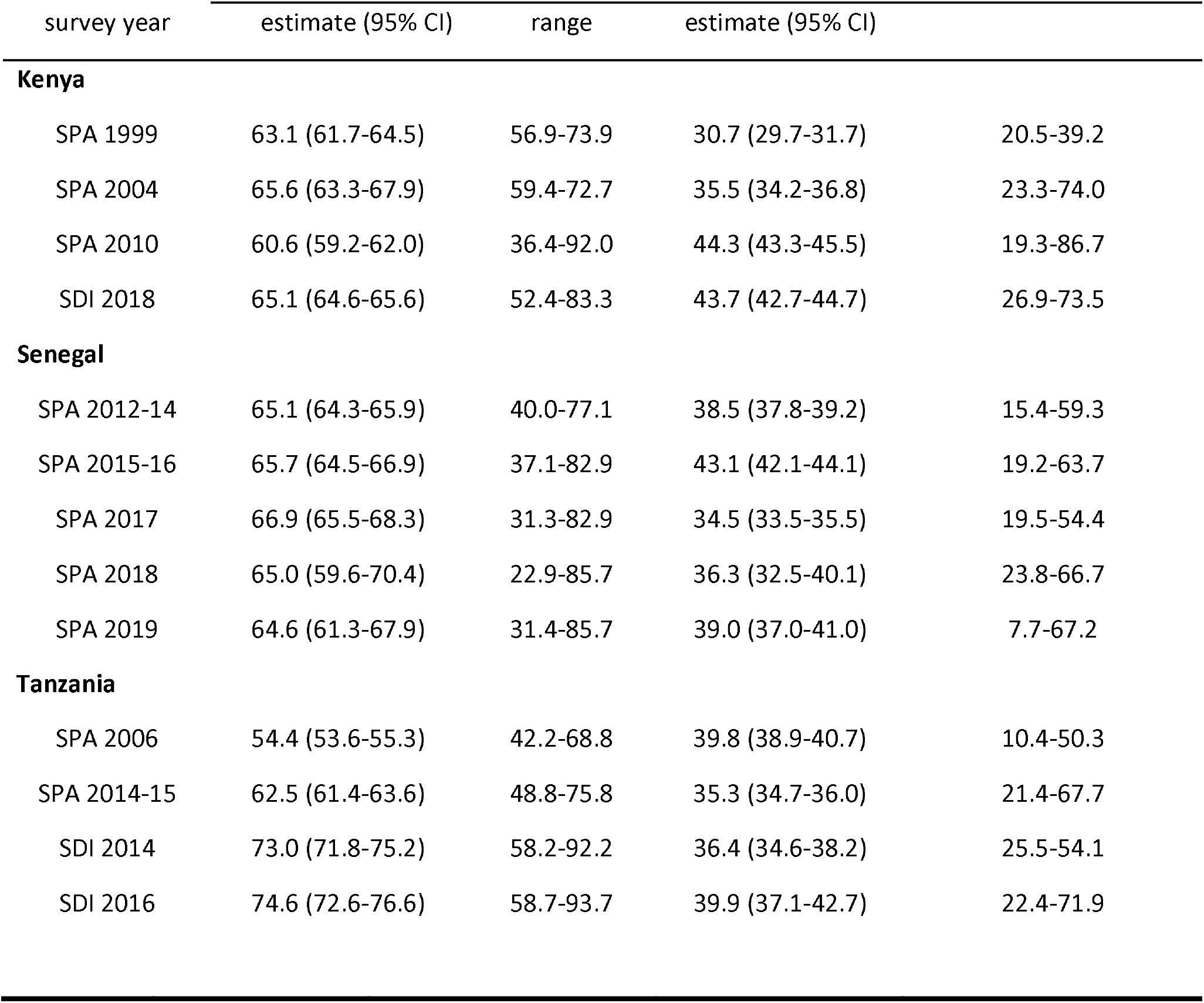
Empirical survey estimates of the national average and the range across subnational areas of the readiness and process quality metrics in Kenya, Senegal, and Tanzania. Estimates presented in this table are empirical survey estimates (as opposed to the modeled estimates presented in what follows). They were calculated separately for each SPA and SDI survey, using their respective survey weights and design variables. The absolute range for each survey was derived by estimating readiness and process quality in each subnational area. Although these surveys were typically not powered to provide reliable estimates at this subnational resolution, we present the absolute range to illustrate the width of subnational inequities within in each country.

*Survey estimates of readiness and process quality metrics*
| Country and | Readiness |  | Process quality |  |
| --- | --- | --- | --- | --- |
|  | Mean survey | Absolute | Mean survey | Absolute range |

**Table 2: Empirical survey estimates of the national average and the range across subnational areas of the readiness and process quality metrics in Kenya, Senegal, and Tanzania.**
| survey year | estimate (95% CI) | range | estimate (95% CI) |  |
| --- | --- | --- | --- | --- |
| **Kenya** |  |  |  |  |
| SPA 1999 | 63.1 (61.7-64.5) | 56.9-73.9 | 30.7 (29.7-31.7) | 20.5-39.2 |
| SPA 2004 | 65.6 (63.3-67.9) | 59.4-72.7 | 35.5 (34.2-36.8) | 23.3-74.0 |
| SPA 2010 | 60.6 (59.2-62.0) | 36.4-92.0 | 44.3 (43.3-45.5) | 19.3-86.7 |
| SDI 2018 | 65.1 (64.6-65.6) | 52.4-83.3 | 43.7 (42.7-44.7) | 26.9-73.5 |
| **Senegal** |  |  |  |  |
| SPA 2012-14 | 65.1 (64.3-65.9) | 40.0-77.1 | 38.5 (37.8-39.2) | 15.4-59.3 |
| SPA 2015-16 | 65.7 (64.5-66.9) | 37.1-82.9 | 43.1 (42.1-44.1) | 19.2-63.7 |
| SPA 2017 | 66.9 (65.5-68.3) | 31.3-82.9 | 34.5 (33.5-35.5) | 19.5-54.4 |
| SPA 2018 | 65.0 (59.6-70.4) | 22.9-85.7 | 36.3 (32.5-40.1) | 23.8-66.7 |
| SPA 2019 | 64.6 (61.3-67.9) | 31.4-85.7 | 39.0 (37.0-41.0) | 7.7-67.2 |
| **Tanzania** |  |  |  |  |
| SPA 2006 | 54.4 (53.6-55.3) | 42.2-68.8 | 39.8 (38.9-40.7) | 10.4-50.3 |
| SPA 2014-15 | 62.5 (61.4-63.6) | 48.8-75.8 | 35.3 (34.7-36.0) | 21.4-67.7 |
| SDI 2014 | 73.0 (71.8-75.2) | 58.2-92.2 | 36.4 (34.6-38.2) | 25.5-54.1 |
| SDI 2016 | 74.6 (72.6-76.6) | 58.7-93.7 | 39.9 (37.1-42.7) | 22.4-71.9 |

Although most SPA and SDI surveys were not powered to directly provide reliable subnational estimates of readiness and process quality, their range suggested potentially important differences in average readiness and process quality between subnational areas in the three countries, over the study period. For instance, process quality spanned from 26.9 to 73.5% in Kenya in 2018, while readiness differed by 40 to 50 points among the departments of Senegal.

### Levels and trends in area-level modelled estimates of readiness and process quality metrics

Marked subnational variations emerged over time for estimated facility readiness and process quality metrics (Figure 1). In Kenya, which had the longest time-series (1999-2018), overall improvements in process quality occurred between 2000 and 2020; this is visualized by shifts for several counties from the upper-left (<50%) to the upper-right quadrant (>50%). However, subnational gains took place amid increasing disparities between counties in both readiness and process quality metrics, as illustrated by the increasing spread of county-level data points each year. Estimated mean readiness ranged from 60.4% (43.1-75.9) to 69.4% (52.9-82.4) in 2000 but from 51.5% (42.4-60.5) to 79.1% (72.1-86.1) in 2020; similarly, estimated process quality spanned from 23.7% (7.8-53.5) to 38.5% (21.1-59.25) in 2000, and 25.8% (16.1-35.6) to 83.3% (74.2-92.4) in 2020. For Senegal, readiness substantially improved in most departments between 2010 and 2020, while process quality stalled. Yet absolute disparities between departments persisted amid overall progress or stagnation; estimated mean readiness ranged from 45.1% (31.6-59.3) to 81.0% (70.4-88.4) and process quality from 26.5% (14.7-42.8) to 61.6% (43.1-77.3), in 2020. Across Tanzanian regions, estimated mean readiness uniformly increased between 2005 and 2020; process quality of care was, however, estimated to stagnate or decline in most regions over the study period.

**Figure 1:**
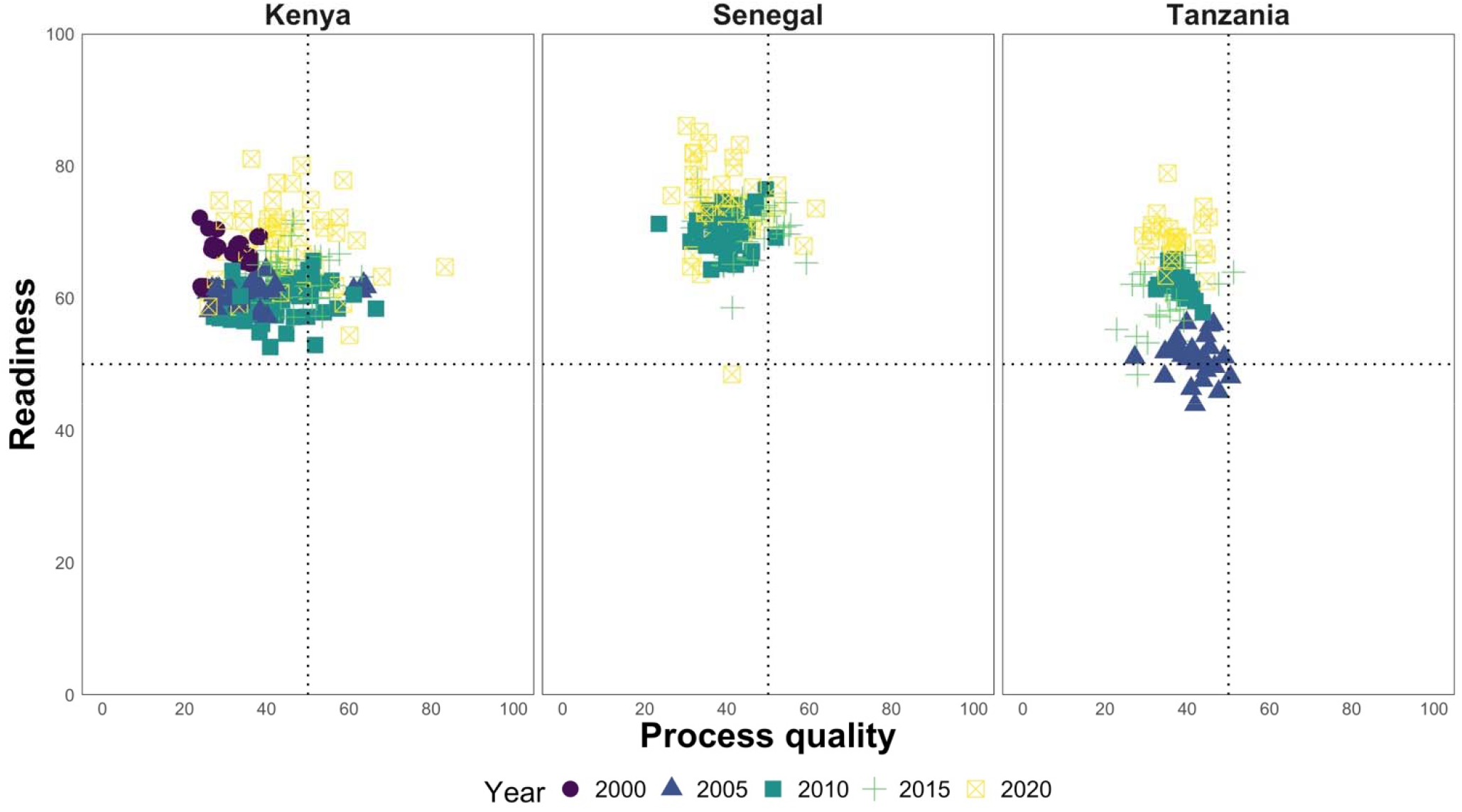
Subnational estimates of readiness and process quality metrics in Kenya, Senegal, and Tanzania in 2000, 2005, 2010, 2015, and 2020. Each point represents a subnational area’s annual estimate of mean process quality (x-axis) and readiness (y-axis). Points in the upper-right quadrant represent areas with average readiness and process quality of care over 50%. The spread of the points horizontally (resp. vertically) represents the variability between subnational areas (points of the same shape and colour) and over time (points of different shapes and colours), in process quality (resp. readiness) across each country.

### Spatiotemporal variations in readiness and process quality metrics in Kenya

In Kenya, readiness of care stagnated overall between 1999 and 2018 (Fig. 2a). Improvements in process quality occurred in most provinces between 2004 and 2010 (Fig. 2b), except for the North-Eastern and Western provinces; in the former process quality stagnated between 2004 and 2010 but improved substantially between 2010 and 2018, while in the latter process quality improved early, but dropped back to levels similar to that of other provinces between 2004 and 2010. Lower readiness and process quality metrics were estimated across counties of the North-Eastern province in 2010 (Fig. 2c), and most counties of the Eastern province. Isiolo and Embu were notable exceptions, as both counties located in the Eastern province had estimated readiness and process quality in the top tercile. Higher readiness and process quality metrics were estimated across most counties of the Nyanza province-Siaya, Kisumu, Migori, Kisii, and Nyamira, along with West Pokot and Narok in the Rift Valley province. These spatial patterns had substantially changed in 2018, when higher readiness and process quality metrics were estimated across counties of the Western province - Turkana, Elgeyo Marakwet, Baringo, and Kilifi in Coast province (Fig. 2d).

**Figure 2:**
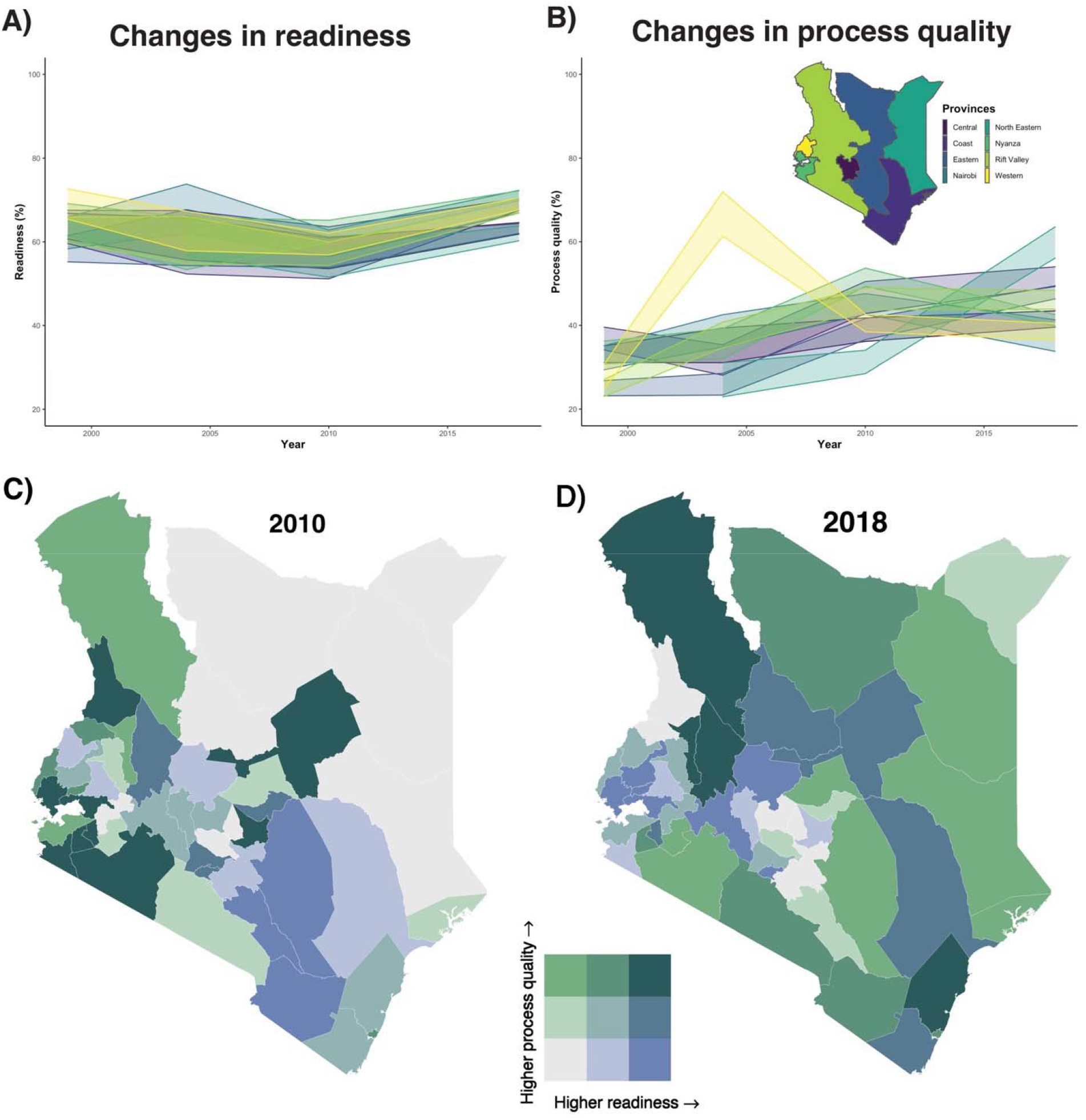
Changes over time in readiness and process quality metrics by subnational areas in Kenya. Predicted mean readiness (A) and process quality (B), by provinces in Kenya between 1999 and 2018. Coloured bands show the 95% posterior prediction intervals for range of the mean estimates across provinces and are colour-coded according to the inset map. **Mean estimated readiness and process quality by counties in Kenya, in 2010 (C) and 2018 (D)**. Mean estimated metrics are split into terciles. These cut-off points indicate the readiness minimum, the 33rd, 66^th^ percentiles, and maximum, which were 52.6%, 57.8%, 60.5%, and 65.6% in 2010, and 57.7%, 64.1%, 69.0%, and 77.8% in 2018. The process quality minimum, 33rd and 66^th^ percentiles, and maximum were 27.1%, 38.2%, 46.7%, and 66.5% in 2010, and 32.8%, 42.0%, 47.2%, and 72.8% in 2018.

### Spatiotemporal variations in the readiness and process quality of care metrics in Senegal

Steady improvements in the readiness of care between 2012 and 2020 in Senegal’s central and northern areas contrasted with stagnation and decline in the southern and western areas, respectively (Fig. 3a). Process quality of care showed very little change, registering around 40% throughout the study period (Fig. 3b). Departments with both higher estimated readiness and process quality of care metrics were clustered in the northern and central parts in 2012: Ranerou Ferlo and Podor (north), Mbacke, Kaffrine, Birkelane and Nioro du Rip (central) had estimated readiness and process quality metrics in the top tercile (Fig. 3c). Conversely, departments with both lower estimated readiness and process quality metrics were concentrated in the western – Pikine – and southern areas of the country - Tambacounda, Goudiry and Bounkiling. By 2020, these regional patterns had been attenuated with several southern departments in the top tercile for both readiness and process quality of care (e.g., Bounkiling, Goudomp, Salemata and Kedougou), and some central departments in the lowest tercile (e.g., Linguere and Louga).

**Figure 3:**
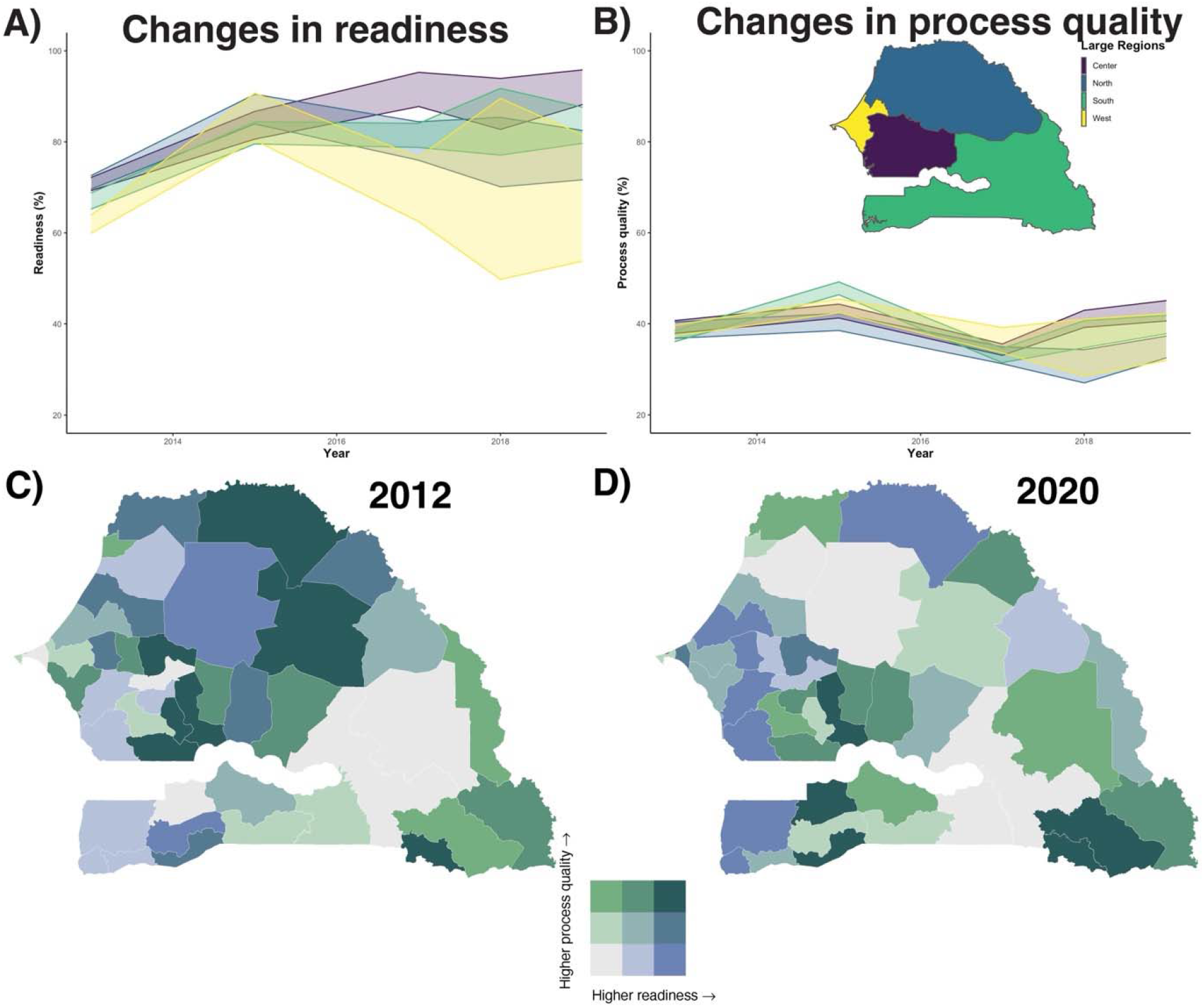
Changes over time in readiness and process quality metrics by subnational areas in Senegal. Predicted mean readiness (A) and process quality (B), by large regions in Senegal between 2012 and 2020. Coloured bands show the 95% posterior prediction intervals for range of the mean estimates across provinces and are colour-coded according to the inset map. **Mean estimated readiness and process quality by departments in Senegal, in 2012 (C) and 2020 (D)**. Mean estimated metrics are split into terciles. These cut-off points indicate the readiness minimum, the 33rd, 66^th^ percentiles, and maximum, which were 53.3%, 66.5%, 71.3%, and 75.4% in 2012, and 46.6%, 73.0%, 76.6%, and 87.1% in 2020. The process quality minimum, 33rd and 66^th^ percentiles, and maximum were 28.3%, 36.5%, 40.6%, and 49.9% in 2012, and 25.1%, 34.8%, 42.1%, and 62.8% in 2020.

### Spatiotemporal variations in readiness and process quality metrics in Tanzania

Readiness of care improved in all large regions of Tanzania between 2006 and 2016, except for a setback in 2015 (Fig. 4a). Process quality stagnated between 2006 and 2014 in the Central, Eastern, and Lake regions, and decreased in the Southern, Southern Highlands, Western, and Zanzibar regions, before increasing everywhere between 2014 and 2016 (Fig. 2b), except in the Northern region, where it decreased. The Southern part of the country was marked by stark contrasts in 2006: Lindi recorded estimated readiness and process quality metrics in the bottom tercile while Mtwara was in the top tercile for both metrics (Fig. 4c). Higher readiness and process quality metrics were otherwise found in the Eastern part of Tanzania and in Zanzibar in 2006, with Morogoro, Unguja North, and Unguja South, in the top tercile. These spatial patterns substantially changed by 2016, when higher readiness and process quality metrics were estimated across regions of the Northern and Central parts of the country – Kagera, Mwanza, Shinyanga, and Singida, and Manyara, respectively (Fig. 4d). Lower readiness and process quality metrics were estimated in Lindi, Tabora, Pemba North, and Pemba South.

**Figure 4:**
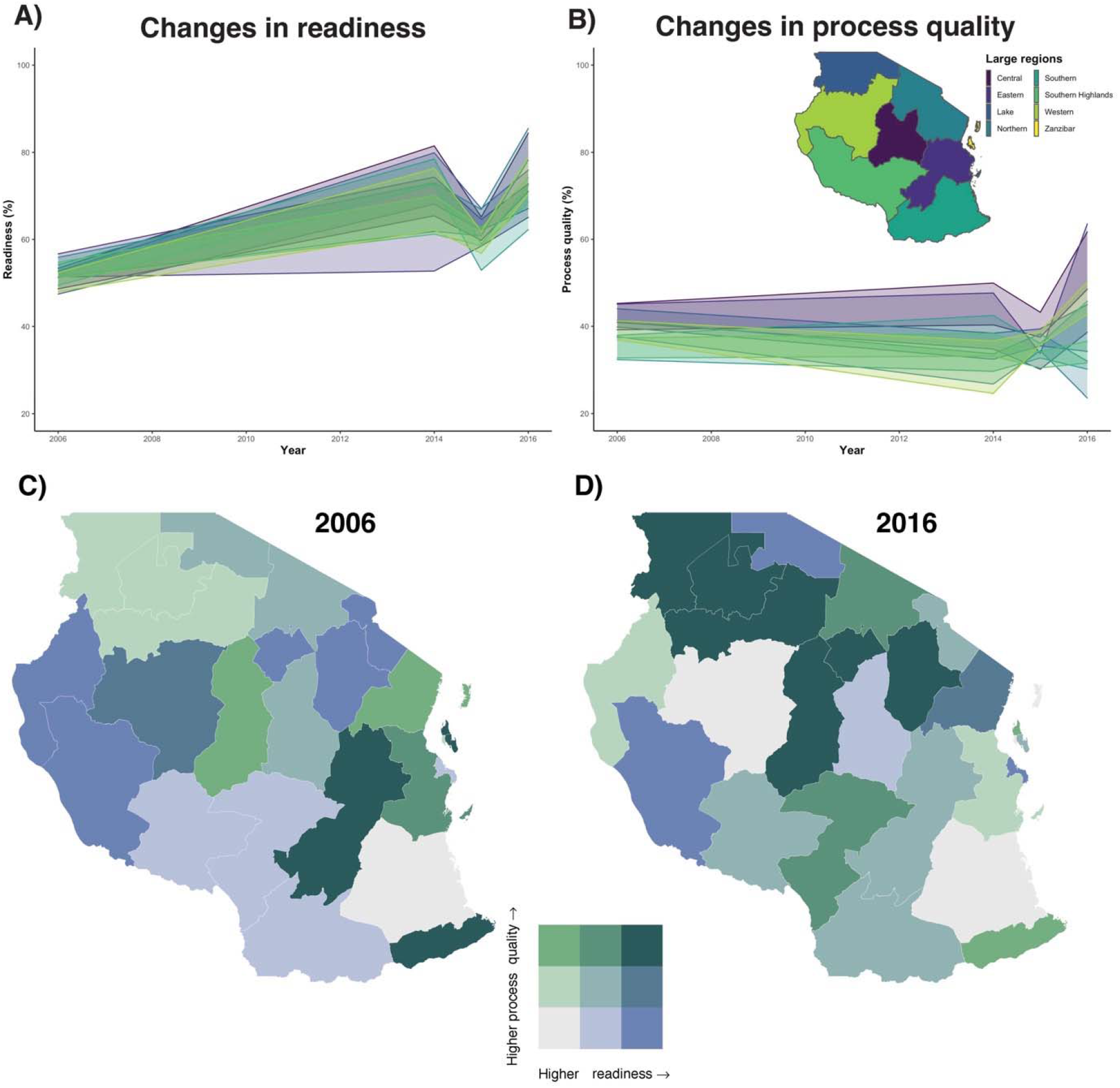
Changes over time in readiness and process quality metrics by subnational areas in Tanzania. Predicted mean readiness (A) and process quality (B), by large regions in Senegal between 2006 and 2016. Coloured bands show the 95% posterior prediction intervals for range of the mean estimates across provinces and are colour-coded according to the inset map. **Mean estimated readiness and process quality by departments in Tanzania, in 2006 (C) and 2016 (D)**. Mean estimated metrics are split into terciles. These cut-off points indicate the readiness minimum, the 33rd, 66^th^ percentiles, and maximum, which were 44.4%, 51.1%, 52.8%, and 57.5% in 2006, and 48.4%, 57.7%, 62.1%, and 66.5% in 2016. The process quality minimum, 33rd and 66^th^ percentiles, and maximum were 25.9%, 37.9%, 43.2%, and 49.3% in 2006, and 22.7%, 31.4%, 37.8% and 51.2% in 2016.

### Comparison of subnational-level model and survey estimates and model validation

Figure 5 compares subnational-level model estimates for the readiness (top) and process quality (bottom) metrics with empirical survey data to which the model was fitted in Senegal, in 2019. Survey estimates fell closely to model mean estimates and within the 95% prediction interval in all but one department for readiness, and two departments for process quality; however, the survey 95% confidence intervals span 0-100% indicating that few to no observations were available in these departments, for that year. In most departments, especially those with few observations, like Birkelane, Koumpentoum and Salemata, the model 95% posterior prediction interval was significantly narrower than the survey 95% confidence interval, highlighting the precision gains that resulted from using a model that borrows information across space and time. We present similar results for the most recent surveys available in Kenya and Tanzania in Supplementary Material 3.3.

**Figure 5:**
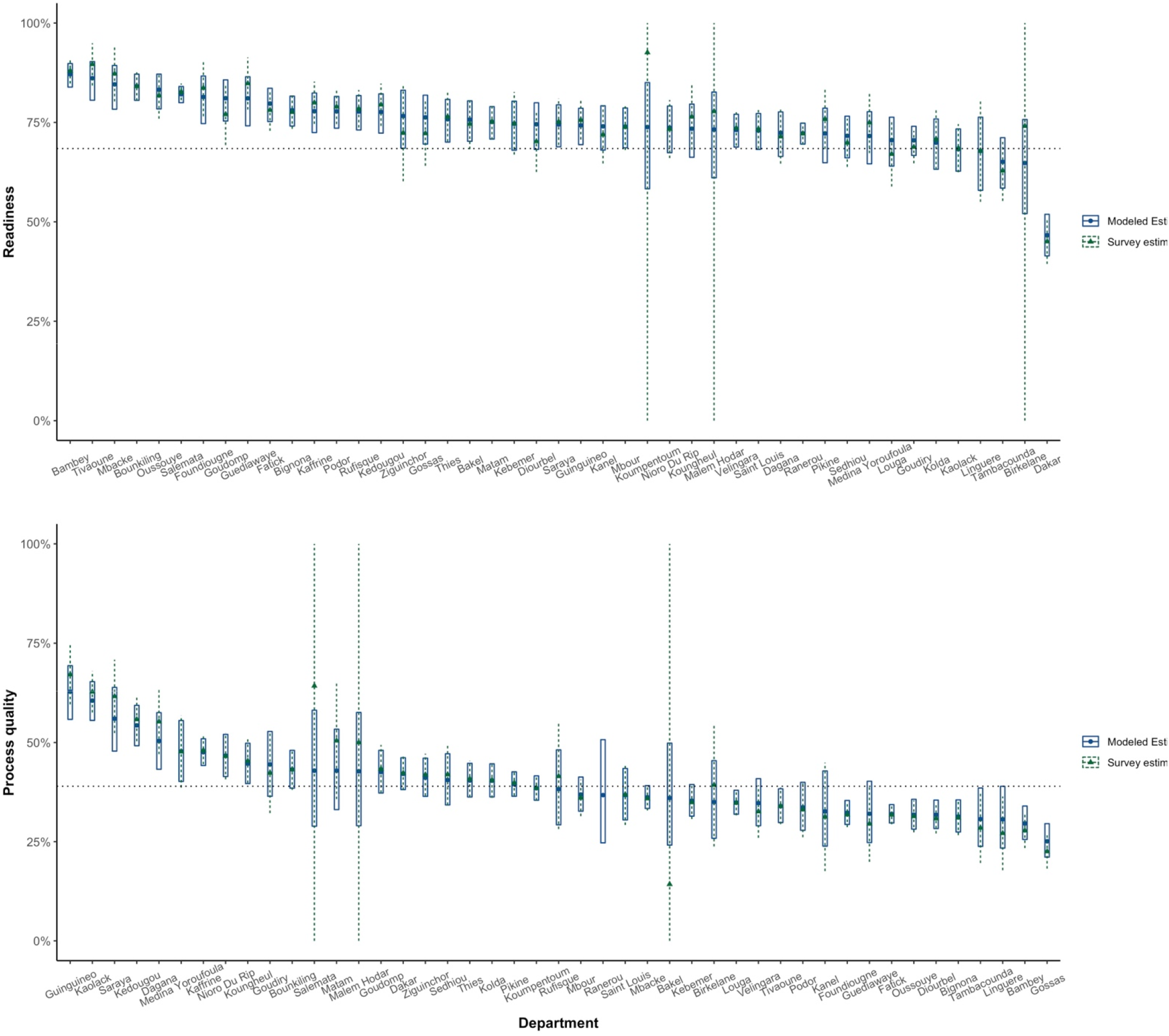
Comparison of department-level survey and model estimates for the readiness and process quality metrics in Senegal, in 2019. This figure compares empirical survey and model estimates, for the most recent year when data was available in Senegal. Blue dots and vertical ranges show model posterior mean estimates, and the 95% posterior prediction intervals. Green triangles and dotted lines indicate direct survey estimates and 95% confidence intervals, derived from SPA 2019. For the readiness metric (top plot), departments are sequenced in decreasing order according to their mean readiness in 2019.

The predictive validity of the models, assessed using hold-one-out area procedure, showed high coverage and low mean squared error (Table 3). Intervals contained the subnational survey estimates over 90% of the time overall, across the three countries. Metric/year/country-specific coverages ranged from 75% to 95%, with notably lower coverage for the readiness metric in Tanzania. The mean difference between the predicted and observed estimate was also highest for the readiness metric in Tanzania, as well as the readiness metric in Senegal. MSE for both metrics in Kenya, and process quality in Senegal and Tanzania were substantially lower, suggesting better model predictive validity for these countries/metrics.

**Table 3:**
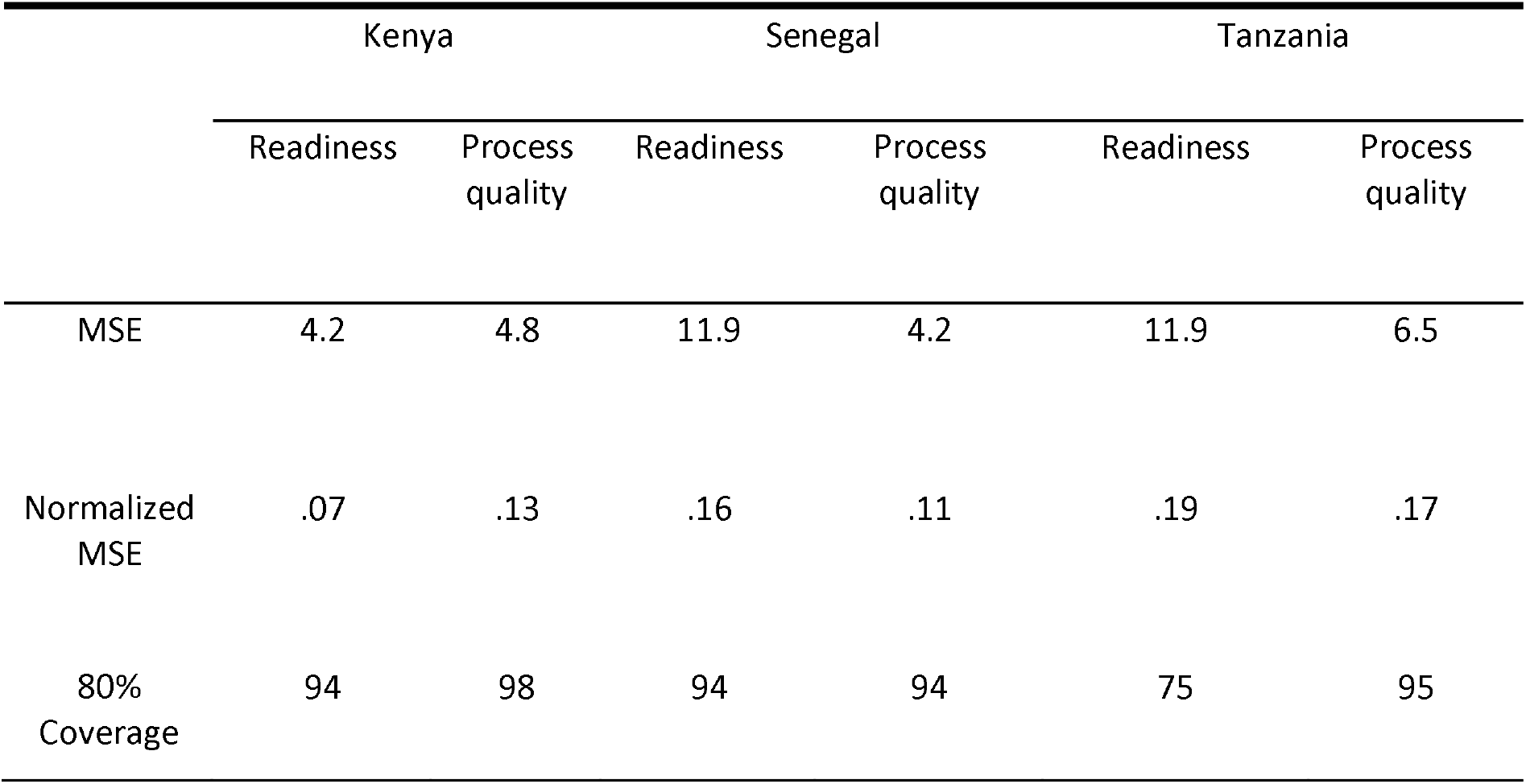
Mean squared error and 80% coverage for hold-out predictions of empirical survey estimates in subnational areas of Kenya, Senegal, and Tanzania. Mean squared errors (MSE) are calculated as the average squared difference between the direct survey estimate for an area and its model prediction, holding-out all the observations for that area. To account for differences in scale between countries and metrics, and facilitate comparison, the normalized MSE is calculated as the MSE divided by the mean survey estimate for that country and metric and 80% coverage are used to assess the models’ predictions. The 80% coverage indicates the frequency at which an area’s survey estimate falls within the model’s 80% prediction intervals for that area, holding-out all the observations for that area while fitting the model.

|  | Kenya |  | Senegal |  | Tanzania |  |
| --- | --- | --- | --- | --- | --- | --- |
|  | Readiness | Process quality | Readiness | Process quality | Readiness | Process quality |
| MSE | 4.2 | 4.8 | 11.9 | 4.2 | 11.9 | 6.5 |
| Normalized MSE | .07 | .13 | .16 | .11 | .19 | .17 |
| 80% Coverage | 94 | 98 | 94 | 94 | 75 | 95 |

## Discussion

Monitoring subnational healthcare quality is critical for identifying and addressing geographic inequities in service provision. With this study, we offer a framework to generate estimates of health facilities’ readiness and process quality of care at a subnational resolution. Our Bayesian hierarchical model supports the incorporation of all available health facility survey data, and accounting for the specific design and uncertainty of each data collection instrument. When recent health facility surveys are not available, our approach leverages spatiotemporal smoothing and auxiliary information from geo-referenced covariates to produce subnational annual estimates of readiness and process quality. Our model-based estimates of readiness and process quality of care over time and at a fine spatial resolution show that progress towards improving facility-based service provision has been uneven within Kenya, Senegal, and Tanzania; for instance, overall progress in readiness of care at the national level in Tanzania masked persistent geographic inequities, while stagnation in this metric in Senegal or Kenya has been associated with enhanced subnational disparities.

Our study expands on previous work on healthcare quality measurement in several ways. First, our study demonstrates the benefits of using a modelling approach alongside direct survey measurements to estimate metrics of readiness and process quality. Several rounds of surveys from the same or from different data collection tools can be analysed jointly, with fixed or random effects accounting for differences between survey instruments. Spatiotemporal smoothing leverages correlation structures in space and time to improve the precision of estimates when data are sparse ^35^, while covariates further improve precision by separating the sources of variability underlying the temporal and geographic distribution of our indicator of interest ^36^. These precision gains allowed us to estimate healthcare quality metrics at a policy-relevant resolution – here, counties (Kenya), departments (Senegal), and regions (Tanzania) – a spatial scale for which surveys were not powered to directly provide reliable estimates. Additionally, in years or regions where no data was collected, our approach imputed data gaps, providing estimates of readiness and quality metrics with associated uncertainty. This feature of our approach provides means to supplement direct estimates derived from surveys that are typically conducted irregularly, with readily available and continuously collected spatially and temporally indexed covariates. Finding covariates relevant to healthcare quality is however challenging; modelled-based estimates are not sufficient to address data gaps but can be useful when the alternative is no estimates at all. We used hold-one-out predictions to test the predictive validity of our models and found high coverages and low mean squared errors. Practically, the complete time-series of these modelled healthcare quality metrics could be used to improve subnational estimates of effective coverage. To date, effective coverage-a key metric to monitor progress towards universal health coverage in LMICs ^37,38^, has been calculated by combining estimates from health facility and household surveys, potentially conducted in different years ^39^; however, this approach can prove problematic if healthcare quality changes rapidly over time ^16,40^. Yearly subnational estimates of healthcare quality metrics mitigate this problem by computing effective coverage from temporally aligned facility and population estimates, with associated uncertainty.

Second, important subnational gaps can be better captured through a more formal space-time modelling framework, rather than limiting analyses to more coarse geographic levels of assessment. For instance, the relative stability in readiness of care in Kenya and Senegal, and in process quality of care in Senegal and Tanzania, at the national level, masked inequitable geographic distributions of healthcare quality metrics in each country. In Kenya and Senegal, the range of estimated readiness of care by counties and departments doubled over the study period. Overall national gains in process quality in Kenya and readiness in Tanzania took place with rising subnational disparities. In Kenya, Senegal, and Tanzania, the progressive devolution of responsibilities, including health service provision, from the national to the local level of government ^23^ might be a driving factor of the diverging trends in healthcare quality between subnational areas; local decision-makers are indeed increasingly in charge of the funding of health facilities (e.g., managing medical supplies, commodities, health workforce) ^41^. Furthermore, access to basic amenities, diagnostic capacities, and essential medicines for child services, remained a major challenge in all three countries: while the availability of basic medical equipment, such as thermometer or child/infant scales, was relatively high across the three countries, only half of the facilities had access to electricity, malaria diagnostic tools, and several tracer drugs, such as amoxicillin, oral rehydration salts, or antimalarial medications, were not available in a quarter to a third of facilities offering curative child services. Given the national priority and efforts, across the three countries, invested in reducing child mortality ^25^, the low levels of tracer drugs for common child infections may be concern for alarm. Moreover, despite IMCI guidelines being a central component of Senegal’s, Kenya’s, and Tanzania’s strategy to reduce child mortality ^42^, we estimated low average adherence to IMCI diagnostic protocols. Estimated average adherence to IMCI diagnostic protocols exceeded 50% for only a few sub-national units across Kenya, Senegal, and Tanzania, in 2020. Recent studies conducted in sub-Saharan Africa similarly reported low levels of compliance with IMCI guidelines ^4,7,12,16,29,40^. For instance, using facility data collected in the Republic of Congo, Cameroon and the Central African Republic, Perales and colleagues found that providers had completed 12 of the 15 IMCI diagnostic protocols in only 8% of sick-child visits ^29^, a recent analysis of the 2014 Ethiopian SPA found that the three main symptoms of child diseases were assessed in 51% of consultations ^12^, and a nine-country analysis of SPA surveys (Haiti, Kenya, Malawi, Namibia, Nepal, Rwanda, Senegal, Tanzania, and Uganda) found that performance of new graduates for sick-child care was low (i.e., an average 0.43 out of 10 recommended actions were observed); all together, these analyses point to a need for strengthening pre-service education and supervision ^43^.

Third, our modelling approach can help identify the optimal frequency and scope of health facility assessments, which to date have been conducted as occasional surveys (Kenya and Tanzania), one-time census (Haiti or Malawi), or continuous yearly survey (Senegal since 2012). Our framework provides a tool to identify the sources of variability in each readiness and process quality metrics, which can help inform data collection efforts around more specific target metrics. For instance, if the target of estimation is a metric displaying strong spatial variability and little temporal variability, a less frequent but more geographically diverse sample may be appropriate. Conversely, a regularly conducted survey with smaller samples would be best suited to estimate metrics displaying substantial temporal but lower spatial variability. A previous study building upon the repeated assessment of the same facilities over two rounds of the SPA survey (2013/2014-2015/2016) in Senegal found greater variations over time in process quality compared to readiness of care metrics ^16^; yet in the present study, we found substantial random temporal variations in both process and readiness of care metrics when considering all facilities assessed in each wave, which may reflect both sampling variability and true changes in the quality metrics.

Our study has several limitations. First, the SPA and SDI exclude non-formal healthcare providers, which can comprise a substantial portion of health service delivery in many LMICs ^44^. In Kenya, Senegal, and Tanzania, it was estimated however that less than 1% of sick children with fever, cough, or diarrhoea, were brought to traditional healers by their caretakers ^45^, suggesting that most sick-child consultations are treated by formal health providers. In Senegal, we also excluded health huts, as the assessment instrument differ for these single-room facilities and did not record elements of process quality of care. However, health huts are often in the front-line in the management of sick children in remote rural areas and studying their referral practices to larger health facilities would be an important undertaking ^46^. Second, we estimated process quality metrics from survey instruments using two different assessment methods (direct observation of consultations vs clinical vignettes), although previous studies have shown that there is a difference between what health providers know and what they do ^47,48^. Nevertheless, in Tanzania, where data collection for SPA and SDI overlapped in 2014, estimates of process quality derived from the two assessment methods led to similar estimates (35.3% and 36.4, respectively). Further harmonization between data collection instruments, as entailed by WHO’s new Harmonized Health Facility Assessment initiative (https://www.who.int/data/data-collection-tools/harmonized-health-facility-assessment/), could enhance the comparability of survey estimates. Third, our composite metrics were calculated as summative measures, which assumes the equal importance of different items to quality, and ultimately to health outcomes. In practice, this may not be the case and items could be assigned weights based on their contribution to health improvements. However, such an approach would require equally large assumptions about the relative importance of each item, and we opted for equal weighting, a common approach in the literature to date ^49,50^. Fourth, while a common technique to improve precision, spatiotemporal smoothing can tend to mitigate true disparities between neighbouring areas or spikes due to true rapid changes from one year to the next. For instance, our modelled estimates may underestimate the true effect on readiness and process quality of care of the health workers, 9-month strike in Senegal in 2018 ^51^. A final limitation of this study is that it focuses on the geographic distribution of readiness and process quality, which can mask important differences within subnational areas between managing authority (private/public) level of care (clinics/hospitals) or type of residence (rural/urban). Stratification on these other important determinants of health service provision, along with subnational area, can be integrated in our modelling approach, but would have resulted in small sample sizes in many subnational areas of the three countries considered in this study. We envision that future work could focus on large health facility assessments to investigate cross-sectional subnational variations by managing authority or levels of care. Other directions include investigating how much variation in the quality of sick-child care is related to large sub-national inequities in under-five mortality ^34,52^.

Strengthening health services is critical to reducing the large burden of deaths amenable to healthcare in LMICs. Standardized measurement of facility capacity, readiness, and quality are necessary for optimally assessing service delivery, and thus leveraging data collected from such assessments are key for better understanding where health service gaps and challenges remain. We offer a statistical framework that addresses methodological gaps for assessing trends and geographic inequities in readiness and process quality of care and supports monitoring countries’ progress toward universal health coverage.

## Data Availability

All data produced in the present study are available upon reasonable request to the authors

## Acknowledgments

We are grateful for Dr. Emelda Okiro for her comments and suggestions, and for Dr Nathaniel Henry and Patrick Liu for conversations on health facility access modelling.

Additionally, we acknowledge that the SPA and SDI surveys used in this article depended upon the dedicated efforts of the many individuals who collected the data and who worked to assure data quality, including health workers, staff of the national statistics organizations; without their efforts, this analysis would not have been possible.

