## Supplementary Methods and Results for "A methodological framework to assess temporal trends and sub-national disparities in healthcare quality metrics using facility surveys, with applications to sick-child care in Kenya, Senegal, and Tanzania" (from another version of this preprint: version 1, 2022-07-19)

This appendix provides supplemental data, figures, and more detailed results for “**A methodological framework to assess temporal trends and sub-national disparities in healthcare quality metrics using facility surveys, with applications to sick-child care in Kenya, Senegal, and Tanzania**”

### List of figures and tables

#### Figures

Figure S1: Comparison of county-level survey and model estimates for the readiness and process quality metrics in Kenya, in 2018.

Figure S2: Comparison of region-level survey and model estimates for the readiness and process quality metrics in Tanzania, in 2016.

#### Tables

Table S1. Characteristics of health facilities sampled in the SPA surveys, in Senegal, Kenya, and Tanzania

Table S2. Characteristics of health facilities sampled in the SDI surveys, in Senegal, Kenya, and Tanzania

Table S3. Covariate data sources

Table S4. List of items used to derive the readiness metric, and differences between the three countries

Table S5. List of diagnostic protocols used to derive the process quality metric

Table S6. Models considered to estimate readiness and process quality metrics over time and space

Table S7. Items availability in Kenya for each round of SPA/SDI survey

Table S8. Items availability in Senegal for each round of SPA survey

Table S9. Items availability in Tanzania for each round of SPA/SDI survey

Table S10. Selected models and variance decomposition, for each readiness and process quality of care metrics in each country. $\sigma_{S_{i}}^{2}$, $\sigma_{\gamma_{t}}^{2}$, $\sigma_{\alpha_{t}}^{2}$, $\sigma_{\delta_{it}}^{2}$, $\sigma_{\upsilon_{t}}^{2}$, and $X_{it}\beta$ columns indicate the percentage of total variance explained by each component.

### Section 1. Supplementary Data

#### Section 1.1. Samples of the SDI and SPA surveys

In Senegal, data were collected as part of a Continuous SPA, which achieved a census of all facilities after five rounds of surveys. Half of the hospitals and health centers were selected in the first (2012-13) and third round (2015) of SPA, while the other half were selected in the second (2014) and fourth round (2016), which created a dependent sampling structure between the first four rounds of the continuous SPA. Random samples of facilities were selected for the SPA conducted in 2017, 2018 and 2019. As the higher levels in the process quality metrics, observed in years 2015 and 2016 compared to 2012-13 and 2014, were not found in the later years, we can hypothesize that the facilities and providers who were observed for the second time in 2015 and 2016, had a better knowledge of SPA interviewers’ assessment criteria, which could have led to an enhanced Hawthorne effect.

|  | |  |  | **Senegal** |  |  |  |  | **Kenya** |  |  | **Tanzania** |
| --- | --- | --- | --- | --- | --- | --- | --- | --- | --- | --- | --- | --- |
| SPA 2013 | | SPA 2014 | SPA 2015 | SPA 2016 | SPA 2017 | SPA 2018 | SPA 2019 | SPA 1999 | SPA 2004 | SPA 2010 | SPA 2006 | SPA 2014 |
| Total number of facilities 364 | | 363 | 375 | 371 | 396 | 339 | 361 | 388 | 440 | 703 | 611 | 1200 |
| Facility type  Hospital | | 35 | 38 | 35 | 35 | 29 | 31 | 32 | 172 | 253 | 128 | 263 |
| Health Centre | | 62 | 65 | 61 | 74 | 62 | 64 | 90 | 51 | 101 | 41 | 380 |
| Clinics | | 266 | 272 | 275 | 287 | 248 | 246 | 256 | 217 | 349 | 437 | 557 |
| Managing authority  Public | 305 | 293 | 299 | 288 | 315 | 270 | 288 | 177 | 175 | 351 | 425 | 783 |
| Private | 59 | 70 | 76 | 83 | 81 | 69 | 73 | 211 | 265 | 352 | 186 | 417 |
| Sick child consultations observed | 1307 | 1212 | 1262 | 1026 | 1064 | 715 | 884 | 623 | 1211 | 2016 | 2559 | 4961 |

**Table S1: Characteristics of health facilities sampled in the SPA surveys, in Senegal, Kenya, and Tanzania**

| **Senegal** | | **Kenya** | | **Tanzania** | | |
| --- | --- | --- | --- | --- | --- | --- |
|  | SDI 2010 | SDI 2012 | SDI 2018 | SDI 2010 | SDI 2014 | SDI 2016 |
| Total number of facilities | 151 | 292 | 3,094 | 175 | 383 | 386 |
| Facility type  Hospital | 0 | 51 | 161 | NA | 27 | 30 |
| Health Centre | 142 | 62 | 484 | NA | 84 | 92 |
| Clinics | 41 | 100 | 2,449 | NA | 272 | 264 |
| Managing authority  Public | 151 | 158 | 1,781 | 175 | 269 | 273 |
| Private | 0 | 134 | 1,313 | 0 | 134 | 127 |
| Vignettes | 150 | 625 | 4,430 | 180 | 570 | 543 |

**Table S2: Characteristics of health facilities sampled in the SDI surveys, in Senegal, Kenya, and Tanzania**

#### Section 1.2. Geolocating health facilities

For SPA survey data, we determined in which regions each facility was located using the GPS coordinates provided by the DHS program and the second-level administrative shapefiles that are publicly available from the Database of Global Administrative Areas (https://gadm.org/). The two most recent SPA surveys in Senegal did not collect GPS coordinates, but DHS provided a linkage file to assign a department to each sampled facility. SDI surveys include the administrative units as a variable directly.

#### Section 1.3. Sampling of sick-child visits and vignettes

In the SPA, the survey teams randomly selected three providers of curative care of sick children among all providers in this service present at the facility the day of the assessment. A maximum of five client consultations for each selected provider was observed. The client weights use the facility sampling weight as its base weight and take into account the total number of clients listed and interviewed within each of the sampling stratum, to calculate the probability of a given consultation to be observed. In the SDI, vignettes were administered to a random sample of health providers (doctors, medical assistants, nurses) among all eligible providers present at the facility the day of the assessment. Providers were randomly sampled among all eligible providers at the facility. The weights here simply represent the percentage probability of selection of a provider within each facility.

#### Section 1.4 Differences between the SPA and SDI surveys

The SPA and SDI surveys’ inventory questionnaire mostly collect the same information, as both surveys use WHO’s SARA framework to assess the availability and readiness of key infrastructure and services in facilities, what we referred to as readiness in this study. However, the differences in the methodology used to assess providers’ knowledge and competence, referred to as process quality in this article, differs significantly. In the SPA survey, interviewers directly observe patient-provider consultations using an observation protocol. In the SDI survey, interviewers act as patients and record providers’ questions, examinations, and recommendations to a hypothetical clinical case, known as vignette. Past studies have shown that compliance to protocol checklists tend to be higher with vignettes than with direct clinical observations. To account for these differences in the assessment of process quality between the SPA and SDI surveys, we used survey-specific effects in our model.

| **Covariate** | **Spatial resolution** | **Temporal resolution** | **Source** |
| --- | --- | --- | --- |
| Total population under five years old | Pixel-level | Annual | WorldPop |
| Travel time to nearest settlement >50,000 inhabitants | Pixel-level | Annual | WorldPop |
| Travel time to nearest health facility | Pixel-level | Annual | WorldPop |
| Health worker density | Administrative unit | Census years | Censuses (derived) |
| Urbanicity | Pixel-level | 2015 | ESA and land cover |
| Night-time lights | Pixel-level | Annual | VIIRS and DMSP (harmonized) |
| Educational attainment | Pixel-level | Annual | IHME |
| Human development index | Pixel-level | Annual | IHME |
| Elevation | Pixel-level | Annual | ArcGIS |

DMSP=Defense Meteorological Satellite Program; ESA=European Space Agency; IHME=Institute for Health Metrics and Evaluation; VIIRS= Visible Infrared Imaging Radiometer Suite

#### Table S3. Covariate data sources

### Section 2. Supplementary Methods

#### Section 2.1 List of tracer items and protocols included in the three countries, to estimate readiness and process quality of care

|  | Included (X) | | |
| --- | --- | --- | --- |
|  | Kenya | Senegal | Tanzania |
| **Readiness** |  |  |  |
| Electricity | X | X |  |
| Improved water | X | X |  |
| Privacy of examination room |  | X |  |
| Improved latrine for client use | X | X |  |
| Communication equipment | X | X | X |
| Computer with email | X | X | X |
| Emergency transport | X | X |  |
| Disposal of sharps  Disposal of medical waste | X  X | X  X | X  X |
| Monthly administrative meetings |  | X |  |
| Quality assurance system |  | X |  |
| System to collect opinion |  | X |  |
| Supervision in the last six months |  | X |  |
| Health workers always available |  | X |  |
| Guidelines for IMCI | X | X | X |
| Child scale | X | X | X |
| Infant scale | X | X | X |
| Thermometer | X | X | X |
| Stethoscope | X | X | X |
| Amoxicillin for children | X | X | X |
| ORS | X | X | X |
| Co-trimoxazole for children | X | X | X |
| Paracetamol | X | X | X |
| Malaria diagnostic capacity | X | X | X |
| Antimalarial medication | X | X | X |

**Table S4: List of tracer items and diagnostic protocols used to derive the readiness metric, and differences between the three countries**

|  | Protocols |
| --- | --- |
| **Process quality of care** |  |
| History taking | Provider asked about cough or difficulty breathing, diarrhoea, fever, inability to drink anything, vomiting, and convulsions |
| Physical examination | Provider took child’s temperature, checked for pallor, looked in child’s ear, counted respiration, checked skin turgor for dehydration, undressed child to examine, weighed the child, pressed both feet to check for oedema |

**Table S5: List of diagnostic protocols used to derive the process quality metric**

#### Section 2.2 Model for sub-national estimation of readiness and process quality of care metrics

The four composite indices of readiness and process quality of care described in table 2 were modelled separately using a small area estimation approach. Specifically, we adapted a previously developed Bayesian framework, which models the direct survey estimates as a function of covariates, and space and time components (Mercer *et al.*, 2015). This Bayesian spatial model allows to estimate time series of the true underlying values of the metrics by smoothing over time and space the direct estimates obtained from multiple surveys, with potentially different designs and associated uncertainty. Specifically, in the first stage, we calculate the Horvitz-Thompson estimator of the metric in area *i*, year *t*, and survey *s*, by using the sampling weight *w* associated to each facility *k*:

${p_{its}}^{HT}= \frac{\sum_{k ⋲ i} w_{k,its}y_{k,its}}{\sum_{k ⋲ i} w_{k,its}}$	with variance $V_{i}^{*}$calculated using standard methods, such as jackknife.

To increase the precision of the design-based estimates of the metric, and to predict the metric in areas or years where no data was collected, we use a hierarchical spatial model inspired by Fay and Herriot classic framework:

$$logit\left( {p_{its}}^{HT} \right)\sim N\left( \theta_{its}, V_{its} \right)$$

$$\theta_{its}= X_{it}\beta+ \gamma_{t}+\alpha_{t}+e_{i}+ S_{i}+ \delta_{it}+ \upsilon_{s}$$

Where $X_{it}\beta$ are area-level covariates for area *i* in year *t* (see table S3), $\gamma_{t}$ are temporal random effects modeled as a first order random walk, $S_{i}$are spatially structured random effects (see section 2.3), $\delta_{it}$ is a space-time interaction, $\alpha_{t}, e_{i}$ and $\upsilon_{s}$ are temporal, spatial and survey zero-mean independent random effects.

#### Section 2.3 Spatial random effects

Area-level random effects were used to model the potential influence of unmeasured features of the local health system. The use of spatially structured random effects reflects our assumption that these unobserved characteristics affecting the availability of quality care are likely to be correlated in space. For instance, remote administrative units might face challenges - in accessing steady drug supply chains for essential medicines and testing materials, or in attracting qualified health workers - that are more like adjacent units than that experienced in the capital city. We evaluated alternative models where area-level random effects were assumed to be independent and identically distributed rather than spatially correlated.

#### Section 2.4 Accounting for the sampling design in Senegal

In Senegal, to account for the changing sampling methods (described in B.1.1), we adopted an analytical approach that includes random effects to account for the rounds of SPA that comprised repeated selection of facilities.

#### Section 2.5 Model selection

Direct estimates and design-based variance estimates were computed using the ***survey*** package in R. We fit the Bayesian hierarchical models using the Integrated Nested Laplace Approximation (Rue *et al.*, 2009) and the R-INLA package. We obtained a subset of all included covariates by checking for multi-collinearity using the variance inflation factor with a threshold of 5. For each indicator and country, we compared the 7 models presented in table S4 consisting of different combinations of the covariates selected with the variance inflation factor (VIF) procedure, and spatio-temporal random effects, using three selection procedures (the deviance information criteria, the Watanabe-Akaike information criteria, and the conditional predictive ordinate). In the absence of consensus on a single criterion, when different criteria pointed to different models, we used a majority rule.

| Model | Formula |
| --- | --- |
| 1 | $\gamma_{t}+\alpha_{t}+e_{i}+ \delta_{it}$ |
| 2 | $\gamma_{t}+\alpha_{t}+ e_{i}+ \delta_{it}+ \upsilon_{s}$ |
| 3 | $X_{it}\beta+ \gamma_{t}+\alpha_{t}+e_{i}+ \delta_{it}$ |
| 4 | $X_{it}\beta+ \gamma_{t}+\alpha_{t}+ \delta_{it}+e_{i}+ \upsilon_{s}$ |
| 5 | $\gamma_{t}+\alpha_{t}+S_{i}+ \delta_{it}$ |
| 6 | $\gamma_{t}+\alpha_{t}+ S_{i}+ \delta_{it}+ \upsilon_{s}$ |
| 7 | $X_{it}\beta+ \gamma_{t}+\alpha_{t}+ S_{i}+ \delta_{it}+ \upsilon_{s}$ |

**Table S6: Models considered to estimate readiness and process quality metrics over time and space**

### Section 3. Supplementary Results

#### Section 3.1 Item availability by country

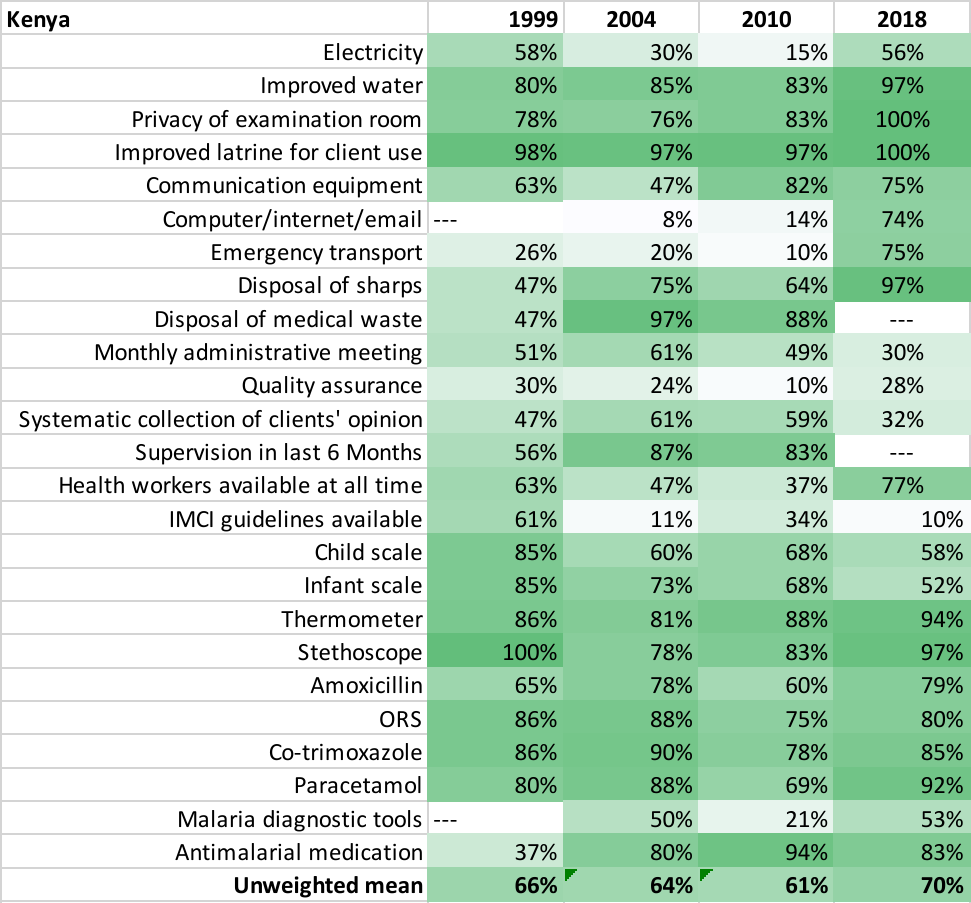

**Table S7: Items availability in Kenya for each round of SPA/SDI survey**. The unweighted mean differs from the readiness metric presented in Table 3, as items are grouped and averaged by domains (infrastructure, facility management, drug availability) before being aggregated as the final readiness metric.

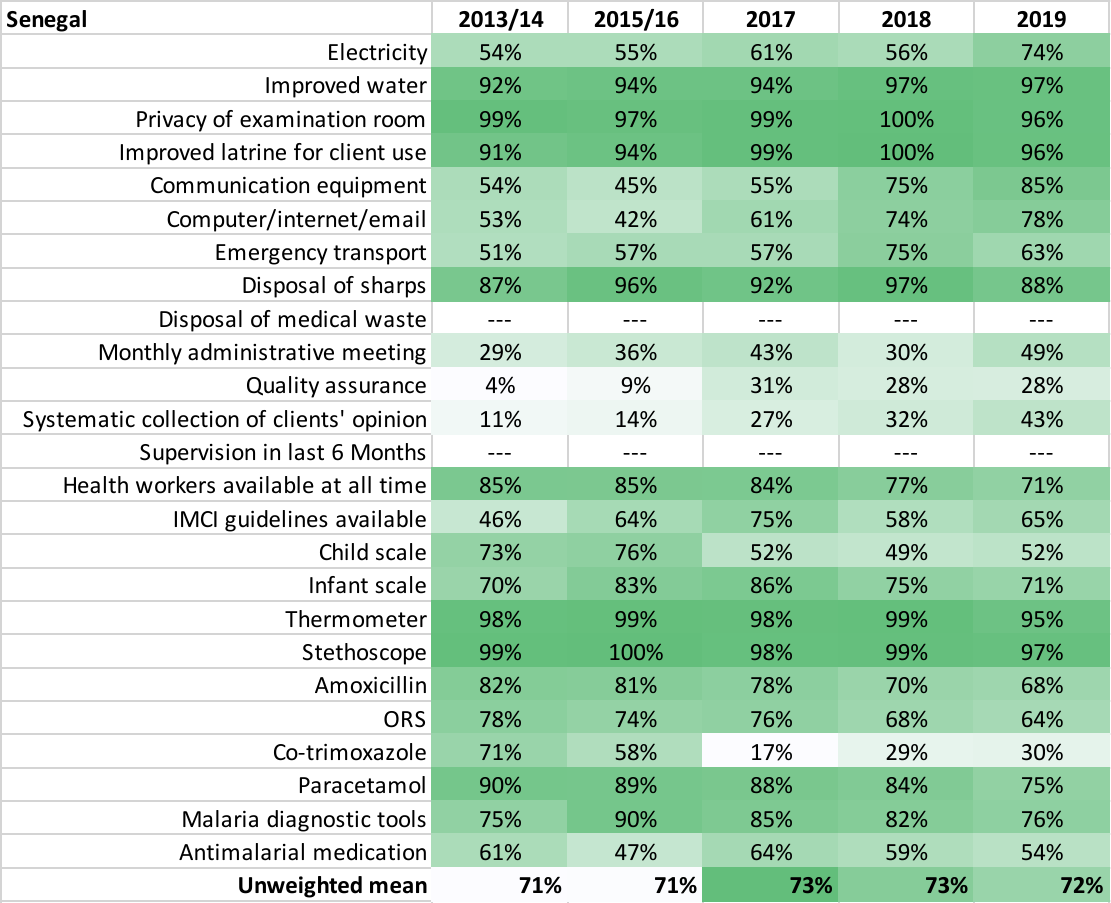

**Table S8: Items availability in Senegal for each round of SPA survey**. The unweighted mean differs from the readiness metric presented in Table 3, as items are grouped and averaged by domains (infrastructure, facility management, drug availability) before being aggregated as the final readiness metric.

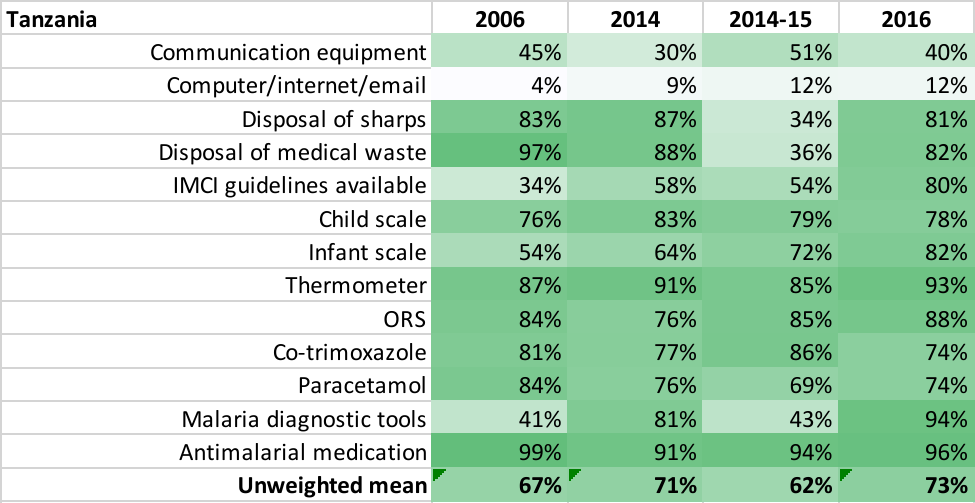

**Table S9: Items availability in Tanzania for each round of SPA/SDI survey**. The unweighted mean differs from the readiness metric presented in Table 3, as items are grouped and averaged by domains (infrastructure, facility management, drug availability) before being aggregated as the final readiness metric.

#### Section 3.2 Decomposition of the sources of variations in the readiness and process quality metrics

|  | **Senegal** | | | | | | | |
| --- | --- | --- | --- | --- | --- | --- | --- | --- |
| Outcome | Model selected | $\sigma_{e_{i}}^{2}$ | $\sigma_{S_{i}}^{2}$ | $\sigma_{\gamma_{t}}^{2}$ | $\sigma_{\alpha_{t}}^{2}$ | $\sigma_{\delta_{it}}^{2}$ | $\sigma_{\upsilon_{s}}^{2}$ | $X_{it}\boldsymbol{\beta}$ |
| Readiness of care | 2 | 27.0 | --- | 2.7 | 1.8 | 64.8 | 3.6 | --- |
| Process quality of care | 5 | --- | 5.1 | 2.3 | 2.9 | 89.7 | --- | --- |
|  | **Kenya** | | | | | | | |
| Outcome | Model selected | $\sigma_{e_{i}}^{2}$ | $\sigma_{S_{i}}^{2}$ | $\sigma_{\gamma_{t}}^{2}$ | $\sigma_{\alpha_{t}}^{2}$ | $\sigma_{\delta_{it}}^{2}$ | $\sigma_{\upsilon_{s}}^{2}$ | $X_{it}\boldsymbol{\beta}$ |
| Readiness of care | 2 | 4.2 | --- | 3.5 | 28.2 | 0 | 64.1 | --- |
| Process quality of care | 7 | --- | 0.7 | 0.1 | 0.7 | 0 | 1.2 | 97.2 |
|  | **Tanzania** | | | | | | | |
| Outcome | Model selected | $\sigma_{e_{i}}^{2}$ | $\sigma_{S_{i}}^{2}$ | $\sigma_{\gamma_{t}}^{2}$ | $\sigma_{\alpha_{t}}^{2}$ | $\sigma_{\delta_{it}}^{2}$ | $\sigma_{\upsilon_{s}}^{2}$ | $X_{it}\boldsymbol{\beta}$ |
| Readiness of care | 3 | 0.6 | --- | 6.3 | 57.3 | 12 | --- | 23.8 |
| Process quality of care | 3 | 0.5 | --- | 1.3 | 0.8 | 53.8 | --- | 43.5 |

**Table S10: Selected models and variance decomposition, for each readiness and process quality of care metrics in each country. The** $\sigma_{e_{i}}^{2}, \sigma_{S_{i}}^{2}$, $\sigma_{\gamma_{t}}^{2}$, $\sigma_{\alpha_{t}}^{2}$, $\sigma_{\delta_{it}}^{2}$, $\sigma_{\upsilon_{t}}^{2}$, and $X_{it}\boldsymbol{\beta}$ **columns indicate the percentage of total variance explained by each component.**

#### Section 3.3 Comparison of subnational-level survey and model estimates for the readiness and process quality metrics in Kenya and Tanzania

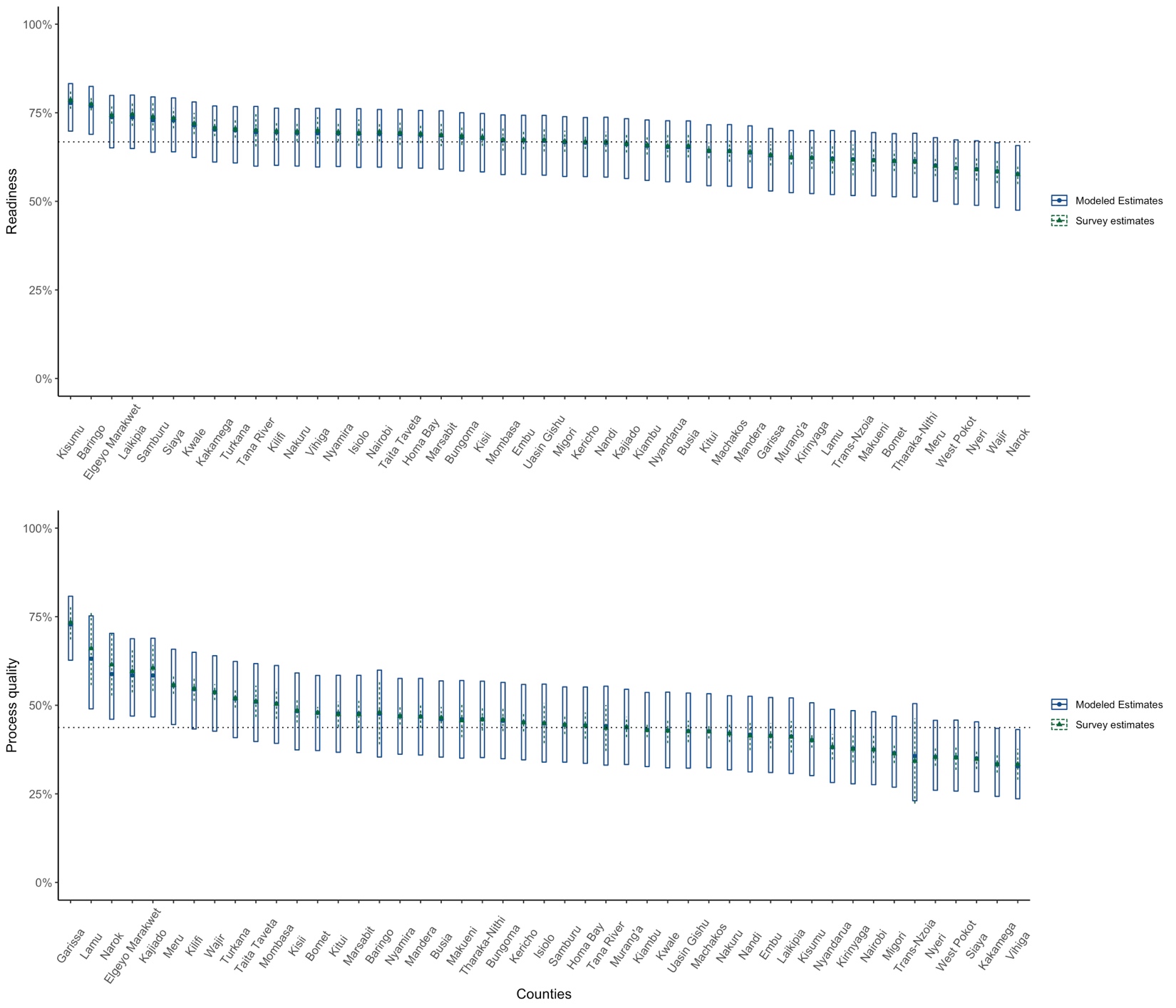

**Figure S1: Comparison of county-level survey and model estimates for the readiness and process quality metrics in Kenya, in 2018.** This figure compares empirical survey and model estimates, for the most recent year when data was available in Kenya. Blue dots and vertical ranges show model posterior mean estimates, and the 95% posterior prediction intervals. Green triangles and dotted lines indicate direct survey estimates and 95% confidence intervals, derived from SDI 2018. For the readiness metric (top plot), counties are sequenced in decreasing order according to their mean readiness in 2018.

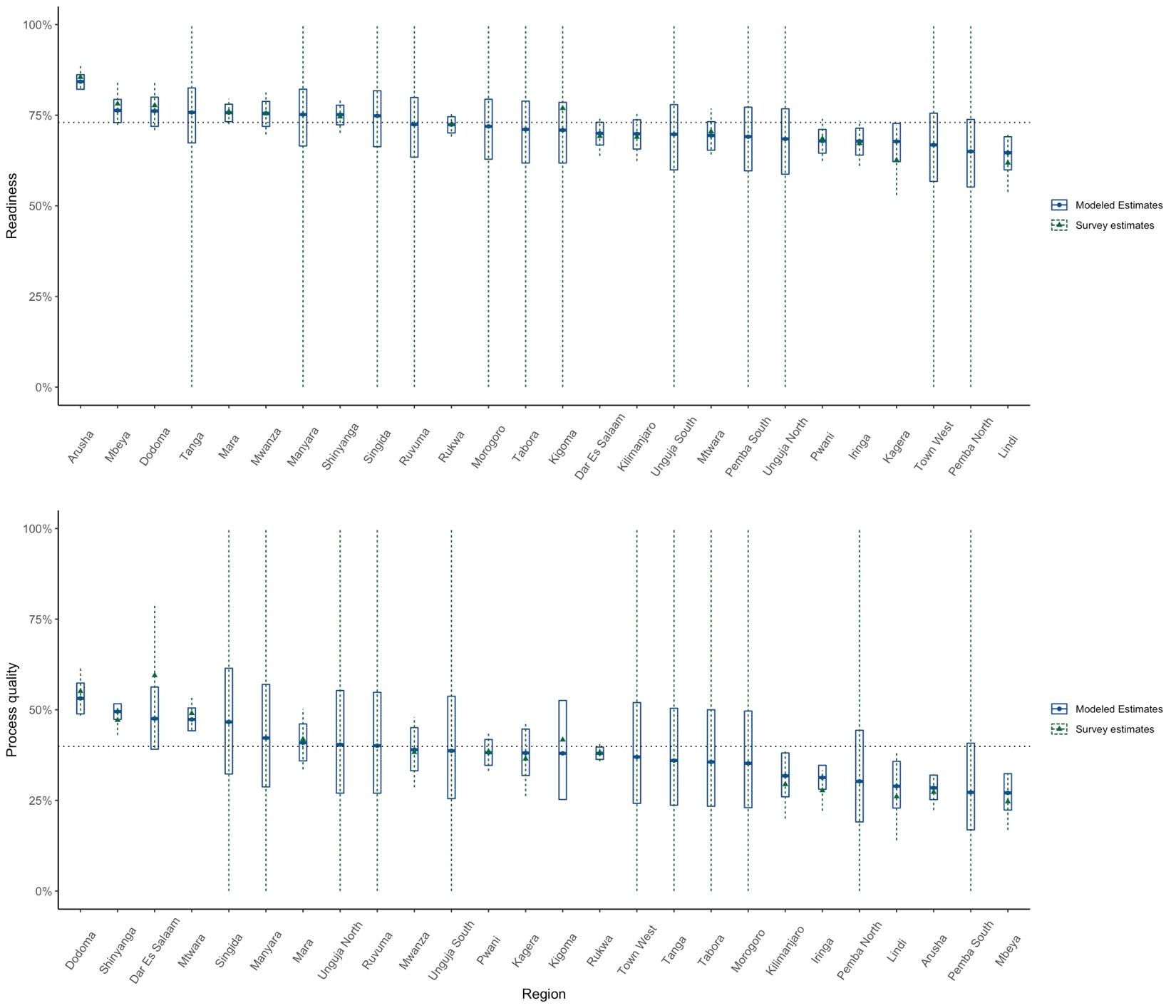

**Figure S2: Comparison of region-level survey and model estimates for the readiness and process quality metrics in Tanzania, in 2016.** This figure compares empirical survey and model estimates, for the most recent year when data was available in Tanzania. Blue dots and vertical ranges show model posterior mean estimates, and the 95% posterior prediction intervals. Green triangles and dotted lines indicate direct survey estimates and 95% confidence intervals, derived from SDI 2018. For the readiness metric (top plot), regions are sequenced in decreasing order according to their mean readiness in 2018.
